# Dementia Risk Reduction in Practice: The Knowledge, Opinions and Perspectives of Australian Healthcare Providers

**DOI:** 10.1101/2020.07.01.20144550

**Authors:** Lidan Zheng, Kali Godbee, Genevieve Z. Steiner, Gail Daylight, Carolyn Ee, Thi Yen Hill, Mark Hohenberg, Nicola T. Lautenschlager, Keith McDonald, Dimity Pond, Kylie Radford, Kaarin J. Anstey, Ruth Peters

## Abstract

**Introduction:** The aim of this paper was to assess Australian primary healthcare providers’ perspectives and knowledge about dementia risk factors and risk reduction.

**Methods:** Primary healthcare providers were recruited through Primary Health Networks across Australia (N = 51). Participants completed an online survey that consisted of fixed-response and free-text components to assess their knowledge, attitudes and current practices relating to dementia risk factors and risk reduction techniques.

**Results:** The survey results showed that over 85% of participants agree that quitting smoking, increasing physical activity, increasing social activity, and treating diabetes can help to reduce the risk of developing dementia. The top suggestions for dementia risk reduction by Australian primary healthcare providers included living a healthy lifestyle (36%), managing cardiovascular risk (17%), and cognitive stimulation (14%). The primary barriers identified for working with patients to reduce dementia risk included low patient motivation and healthcare system level limitations. The most common recommendations were increasing resources and improving dementia awareness and messaging.

**Conclusions:** Collaborative efforts between researchers, media, clinicians, and policy makers are likely needed to support the uptake of risk reduction activities into primary care settings.

## Introduction

The prevalence of people with dementia in the population is rising. In Australia, almost one in 10 people over 65 have dementia ^1^. A range of vascular conditions and lifestyle factors have been linked to increased dementia risk including hypertension, diabetes, stroke, mid-life dyslipidaemia, smoking, and physical inactivity among others (See Table 1 for a list of modifiable risk factors for dementia). Interventions targeting these risk factors have been shown to have the potential to reduce the risk of developing dementia or slow cognitive decline ^2-6^. As many of the risk factors for dementia are also risk factors for other chronic diseases (e.g., cancer, heart disease, diabetes, and stroke ^7^), dementia risk reduction has the potential to be economical and beneficial on multiple fronts.

**Table 1.** List of modifiable risk factors for dementia. Data summarised from a recent systematic review of meta-analyses that evaluated risk factors for dementia ^15^.

| Medical | Cardiovascular | Lifestyle | Pharmacological | Other |
| --- | --- | --- | --- | --- |
| • Body mass index<br>• Depression<br>• Hormones<br>• Sleep<br>• Traumatic brain injury<br>• Arthritis<br>• Cancer<br>• Anxiety<br>• Hearing loss<br>• Inflammatory markers<br>• Motor function<br>• Renal disease | • Diabetes<br>• Cholesterol<br>• Atrial fibrillation<br>• Hyper/hypotension<br>• Stroke<br>• Carotid atherosclerosis<br>• Metabolic syndrome<br>• Homocysteine<br>• Peripheral artery disease<br>• Serum uric acid | • Diet<br>• Physical activity<br>• Smoking<br>• Alcohol<br>• Cognitive engagement<br>• Social engagement<br>• Stress | • Anti-hypertensives<br>• Statins<br>• Anti-inflammatories<br>• Hormone replacement therapy<br>• Antacids<br>• Benzodiazepines<br>• Insulin sensitisers | • Education<br>• Bilingualism<br>• Pesticides |

Primary healthcare providers see a wide range of medical conditions and have relatively frequent contact with a large proportion of the population. The most recent Australian Institute Health and Welfare survey of primary healthcare in Australia showed that more than 80% of the Australian population visit a general practitioner (GP) annually ^8^. Primary healthcare providers are therefore well placed to provide dementia risk reduction advice. However, it is unclear the extent of knowledge that primary healthcare professionals have about dementia risk factors and risk reduction techniques.

Previous research that focused on exploring knowledge about dementia more broadly has found that Australian primary healthcare professionals do not feel confident about their dementia knowledge. Millard et al. (2011) conducted a survey with GPs and nurses (N = 153) and found that only 59% of their sample had received dementia training, and 21% of their sample thought their dementia knowledge was adequate ^9^. However, no studies to date have examined Australian primary healthcare provider knowledge on dementia risk reduction specifically.

This paper explored what primary healthcare professionals currently know about dementia risk reduction. Specifically, it i) examined Australian primary healthcare provider knowledge about dementia risk factors, and ii) identified the barriers and enablers or recommendations to support the adoption of dementia risk reduction activities in primary care.

## Methods

An online survey was conducted with primary healthcare providers within Australia to assess their knowledge of dementia risk factors. The survey was distributed through email, primary health network newsletters, face-to-face direct contacts, social media and flyers at professional development events from November 2018 to November 2019. Inclusion criteria included working in a primary healthcare role (e.g., GP, practice nurse, or allied health professional) in Australia. The survey consisted of several components including:

1. Questions about the characteristics of the participant and their practices.
2. Free-text and fixed-response questions examining factual dementia risk reduction knowledge. Specifically, the free-text component required participants to provide up to five free text responses to the question “What do you think a person can do to help reduce the risk of developing dementia?”. The fixed-response component asked participants to rate on a 5 point Likert scale from “strongly agree” to “strongly disagree” whether or not they felt that there was sufficient evidence for a range of dementia risk reduction techniques (e.g., “Learning a new language”, “Lowering blood pressure”).
3. Free text components on barriers and enablers to working with patients to reduce dementia risk. Specifically, participants were asked to provide free-text answers to the following two questions: (1) “What are the biggest barriers to working with your patients to reduce dementia risk?” and (2) “What helps (or would help) you to work with patients to reduce dementia risk?”.

Statistical analysis of responses was completed using SPSS v25 ^10^ and free text was analysed using inductively coded thematic analysis. Mann-Whitney U and chi-squared tests were used for group comparisons. This study was approved by the UNSW HREC (HC3120).

## Results

A total of 51 participants completed the survey. The respondents were primarily female (N = 44). The average age of respondents was 58.98 years (SD = 11.93). Of the respondents who completed the survey, 20 were GPs and 20 were practice nurses (PNs) while the rest were either registrars or other health workers. The majority (N = 28) had been practicing for less than 10 years. There was representation from every state except from the Northern Territory and a relatively even distribution of primary practice locations across high, medium, and low SES regions. See Table 2.

**Table 2.**
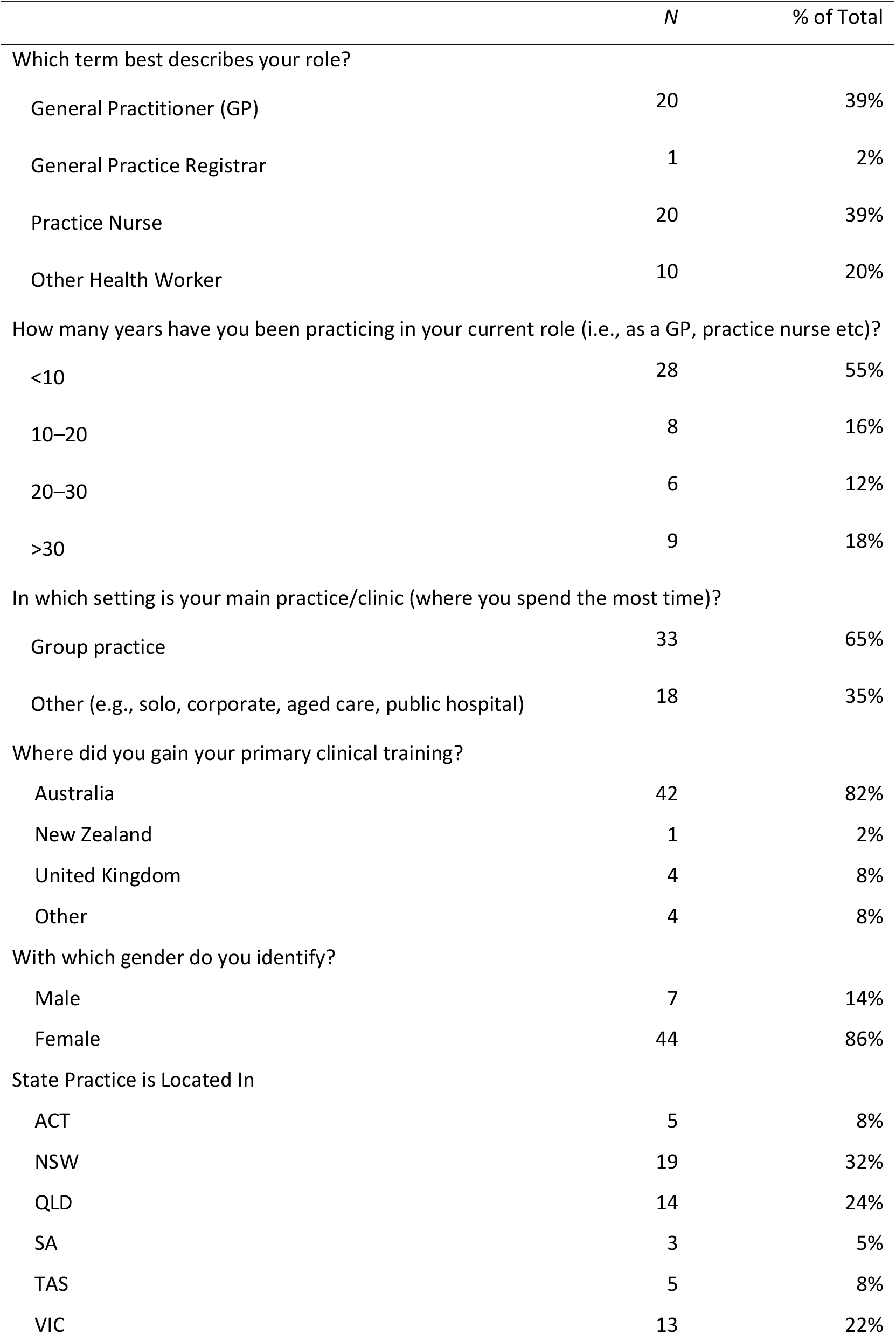
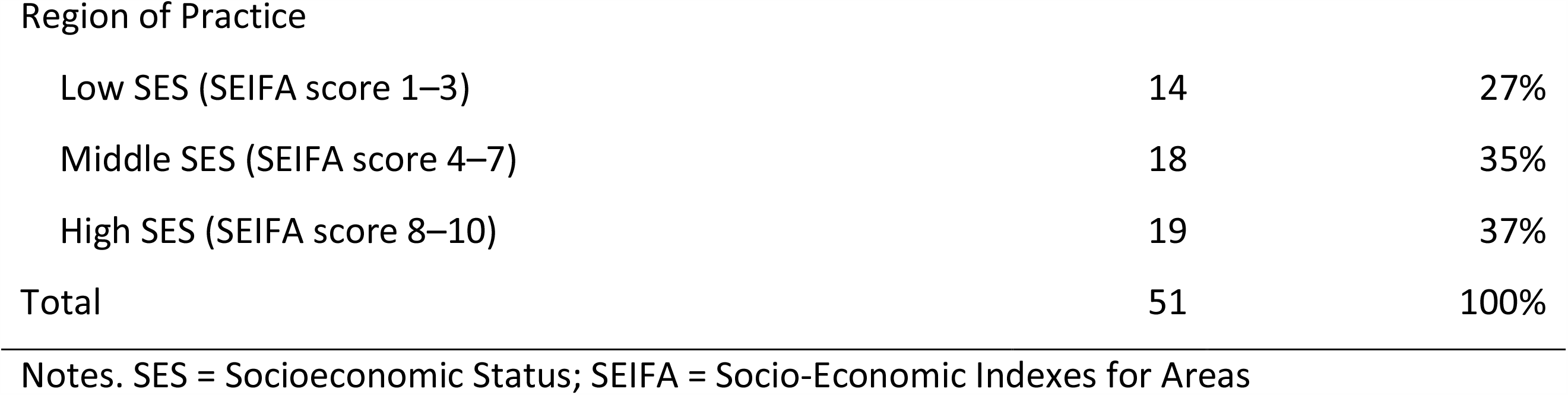
Primary Healthcare Provider Demographics and Practice Characteristics

|  | *N* | % of Total |
| --- | --- | --- |
| Which term best describes your role? |  |  |
| General Practitioner (GP) | 20 | 39% |
| General Practice Registrar | 1 | 2% |
| Practice Nurse | 20 | 39% |
| Other Health Worker | 10 | 20% |
| How many years have you been practicing in your current role (i.e., as a GP, practice nurse etc)? |  |  |
| <10 | 28 | 55% |
| 10–20 | 8 | 16% |
| 20–30 | 6 | 12% |
| >30 | 9 | 18% |
| In which setting is your main practice/clinic (where you spend the most time)? |  |  |
| Group practice | 33 | 65% |
| Other (e.g., solo, corporate, aged care, public hospital) | 18 | 35% |
| Where did you gain your primary clinical training? |  |  |
| Australia | 42 | 82% |
| New Zealand | 1 | 2% |
| United Kingdom | 4 | 8% |
| Other | 4 | 8% |
| With which gender do you identify? |  |  |
| Male | 7 | 14% |
| Female | 44 | 86% |
| State Practice is Located In |  |  |
| ACT | 5 | 8% |
| NSW | 19 | 32% |
| QLD | 14 | 24% |
| SA | 3 | 5% |
| TAS | 5 | 8% |
| VIC | 13 | 22% |
| Region of Practice |  |  |
| Low SES (SEIFA score 1–3) | 14 | 27% |
| Middle SES (SEIFA score 4–7) | 18 | 35% |
| High SES (SEIFA score 8–10) | 19 | 37% |
| Total | 51 | 100% |

Notes. SES = Socioeconomic Status; SEIFA = Socio-Economic Indexes for Areas

### Dementia prevention and risk reduction

The majority of respondents agreed that it is possible to reduce the risk of a person developing dementia (N = 34, 67%). Of the current sample, 34% of participants (N = 17) said that they had received training on dementia risk factors and dementia prevention, and 25% of participants (N = 13) had written information about lowering the risk of dementia available for patients at their practice/clinic. A little over half (N = 29, 57%) said that they had visited a ‘brain health’ website in the past 12 months. On a scale of 0 (“Insufficient”) to 100 (“Sufficient”) knowledge of dementia risk factors, on average, participants considered that the amount of knowledge they had about dementia risk factors around 57%. See Table 3.

**Table 3.** Responses to Questions on Dementia Risk Factors and Risk Reduction

|  | Total<br>(N = 51) | PN<br>(N = 20) | GP<br>(N = 20) | <10 Years<br>Practice<br>(N = 28) | >10 Years<br>Practice<br>(N = 23) |
| --- | --- | --- | --- | --- | --- |
| It is possible to reduce the risk of a person developing dementia |  |  |  |  |  |
| Strongly disagree | 4 (8%) | 2 (10%) | 1 (5%) | 1 (4%) | 3 (13%) |
| Somewhat disagree | 6 (12%) | 3 (15%) | 2 (10%) | 4 (14%) | 2 (9%) |
| Neither agree nor disagree | 7 (14%) | 3 (15%) | 2 (10%) | 4 (14%) | 3 (13%) |
| Somewhat agree | 29 (57%) | 11 (55%) | 13 (65%) | 16 (57%) | 13 (57%) |
| Strongly agree | 5 (10%) | 1 (5%) | 2 (10%) | 3 (11%) | 2 (9%) |
| Have you received training on dementia risk factors and dementia prevention? |  |  |  |  |  |
| Yes | 17 (34%) | 6 (30%) | 4 (20%) | 9 (32%) | 8 (35%) |
| No | 31 (62%) | 12 (60%) | 16 (80%) | 19 (68%) | 13 (56%) |
| I'm not sure | 2 (4%) | 2 (10%) | 0 (0%) | 0 (0%) | 2 (9%) |
| Do you feel that you have sufficient knowledge of dementia risk factors? (0 = Insufficient, 100 = Sufficient). I consider my knowledge: |  |  |  |  |  |
| Mean (SD) | 56.54<br>(28.77) | 37.20<br>(28.80) | 72.82<br>(20.29) | 52.11<br>(30.58) | 63.65<br>(25.69) |
| Does your practice/clinic have any written information available for patients about lowering the risk of dementia? |  |  |  |  |  |
| Yes | 13 (25%) | 2 (10%) | 6 (30%) | 5 (18%) | 8 (35%) |
| No | 24 (47%) | 9 (45%) | 10 (50%) | 14 (50%) | 10 (44%) |
| I'm not sure | 14 (27%) | 9 (45%) | 4 (20%) | 9 (32%) | 5 (22%) |
| Have you visited the Dementia Australia or a similar 'brain health' website in the last 12 months? |  |  |  |  |  |
| Yes | 29 (57%) | 10 (50%) | 12 (60%) | 16 (57%) | 13 (57%) |
| No | 21 (41%) | 10 (50%) | 7 (35%) | 11 (39%) | 10 (43%) |
| I'm not sure | 1 (2%) | 0 (0%) | 1 (5%) | 1 (4%) | 0 (0%) |

Note. Unless specified otherwise, responses are presented as N (%). N = total number of people who chose this response, % = proportion of participants who chose this response.

### Comparisons between practice nurses and general practitioners

PNs were much less confident in their knowledge about dementia risk factors compared to GPs (Mann–Whitney U test: Z = −3.371, p = <.001). However, there were no differences in whether or not they had received training in dementia risk factors and dementia prevention (chi-squared = 2.971, p = .226). There was also no statistical difference between the proportion of GPs and PNs who believed it was possible to reduce the risk of a person developing dementia (Mann–Whitney U test: Z = −1.087, p = .341).

### Relationship to years of practice

There was no relationship between years of practice and beliefs about whether it is possible to reduce the risk of a person developing dementia (Mann–Whitney U test: Z = −.410, p = .682), confidence in knowledge about dementia risk factors (Mann–Whitney U test: Z = −1.26, p = .208), or training on dementia risk factors and risk reduction (chi-squared = 2.962, p = .227).

### Assessment of factual knowledge about dementia risk factors and risk reduction

In the two sections which assessed primary healthcare provider knowledge about dementia risk factors and risk reduction, a total of 242 responses were received for the free-text component (see Appendix 1 for a comprehensive list of responses); 36% (N = 86) of these related to the theme of living a healthy lifestyle (e.g., exercise, healthy diet), 17% (N = 40) to managing cardiovascular disease or decreasing cardiovascular risk, 14% (N = 35) to cognitive stimulation, 13% (N = 31) to reducing smoking and alcohol use, and 9% (N = 21) to increasing social activity. The other 11% (N = 26) were spread out among the themes of mental wellbeing, getting adequate sleep, managing hearing loss, managing medication use and preventing head injury.

A fixed-response section assessed the participant’s awareness of the evidence base by asking healthcare providers’ to rate whether evidence was sufficient for various activities, lifestyle changes or clinical interventions to reduce dementia risk. See Table 4 for a list of responses. The top five interventions that primary healthcare providers perceived as having sufficient evidence for were: quitting smoking (95% somewhat or strongly agree), increasing physical activity (94%), increasing social activity (90%), treating diabetes (86%) and lowering cholesterol (84%). The five interventions perceived as not having sufficient evidence for were: living near powerlines (55% somewhat or strongly disagree), including coconut oil into their diet (55%), avoiding living near busy roads (50%), including turmeric in their diet (48%) and including *Ginkgo biloba* in their diet (43%). The responses, along with the level of evidence for each risk factor according to the literature is reported in Table 4.

**Table 4.**
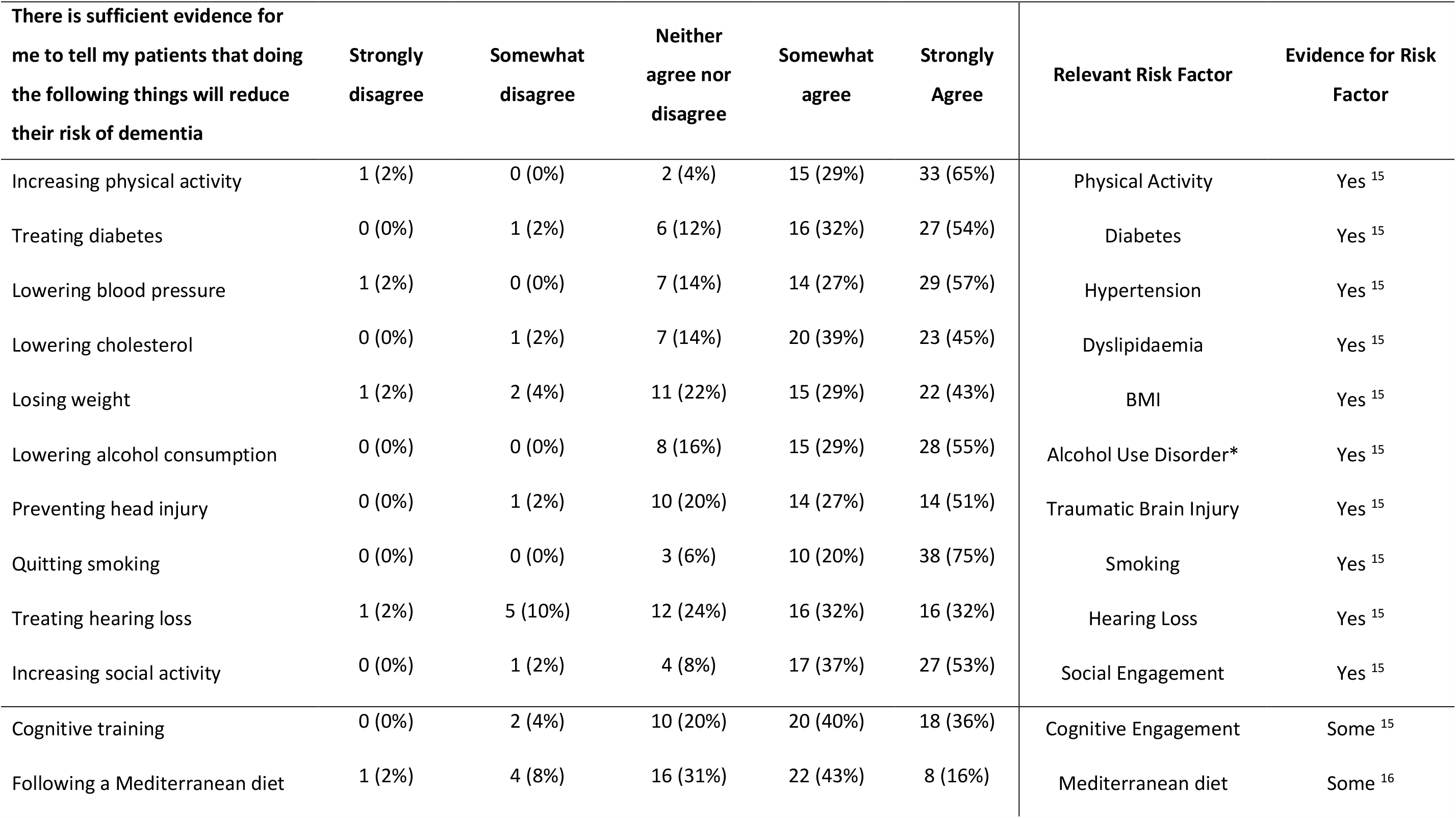
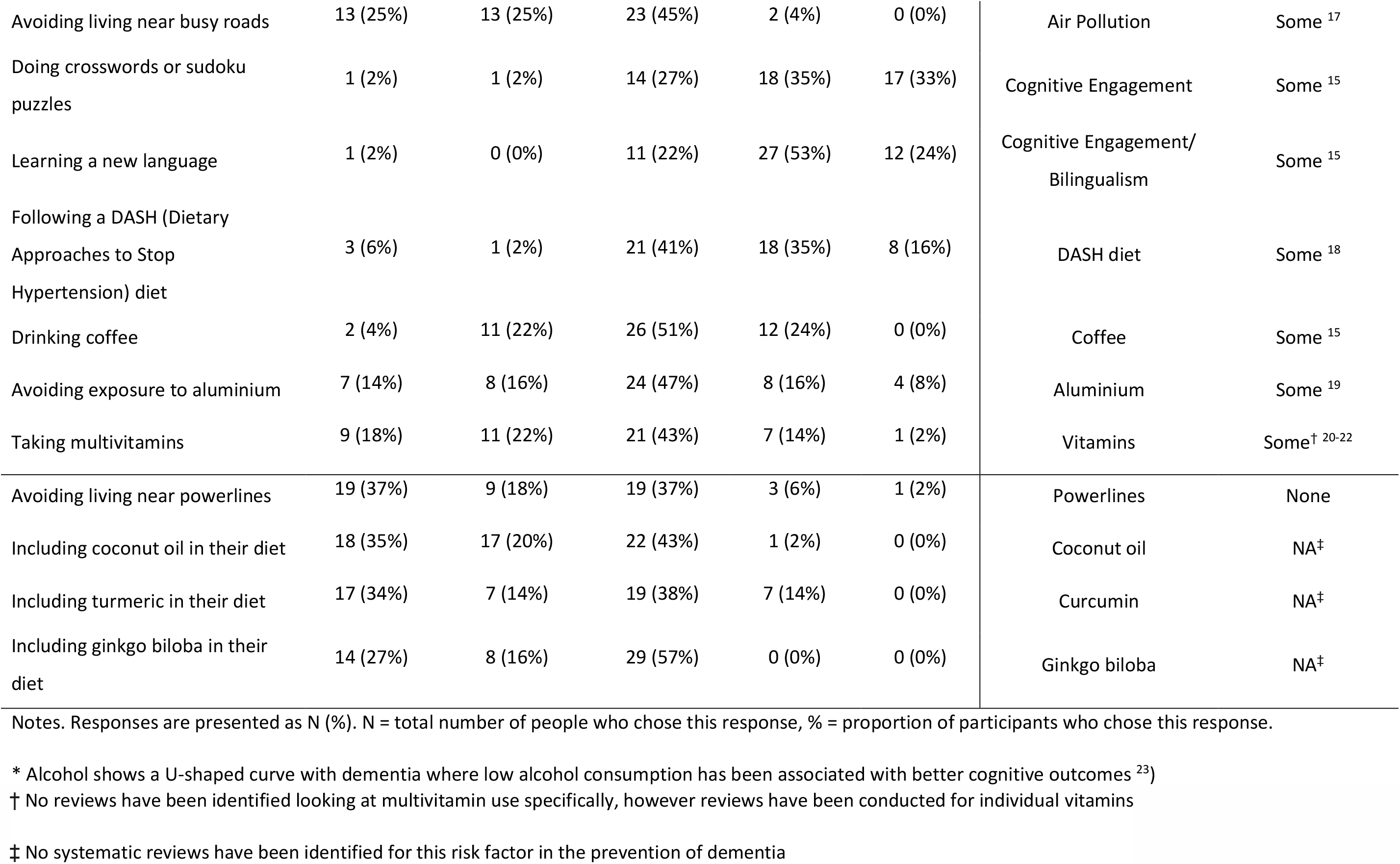
Responses to whether or not there was sufficient evidence for the treatment of dementia risk factors. Ranked in order of level of evidence for risk factor.

| **There is sufficient evidence for me to tell my patients that doing the following things will reduce their risk of dementia** | **Strongly disagree** | **Somewhat disagree** | **Neither agree nor disagree** | **Somewhat agree** | **Strongly Agree** | **Relevant Risk Factor** | **Evidence for Risk Factor** |
| --- | --- | --- | --- | --- | --- | --- | --- |
| Increasing physical activity | 1 (2%) | 0 (0%) | 2 (4%) | 15 (29%) | 33 (65%) | Physical Activity | Yes <sup>15</sup> |
| Treating diabetes | 0 (0%) | 1 (2%) | 6 (12%) | 16 (32%) | 27 (54%) | Diabetes | Yes <sup>15</sup> |
| Lowering blood pressure | 1 (2%) | 0 (0%) | 7 (14%) | 14 (27%) | 29 (57%) | Hypertension | Yes <sup>15</sup> |
| Lowering cholesterol | 0 (0%) | 1 (2%) | 7 (14%) | 20 (39%) | 23 (45%) | Dyslipidaemia | Yes <sup>15</sup> |
| Losing weight | 1 (2%) | 2 (4%) | 11 (22%) | 15 (29%) | 22 (43%) | BMI | Yes <sup>15</sup> |
| Lowering alcohol consumption | 0 (0%) | 0 (0%) | 8 (16%) | 15 (29%) | 28 (55%) | Alcohol Use Disorder* | Yes <sup>15</sup> |
| Preventing head injury | 0 (0%) | 1 (2%) | 10 (20%) | 14 (27%) | 14 (51%) | Traumatic Brain Injury | Yes <sup>15</sup> |
| Quitting smoking | 0 (0%) | 0 (0%) | 3 (6%) | 10 (20%) | 38 (75%) | Smoking | Yes <sup>15</sup> |
| Treating hearing loss | 1 (2%) | 5 (10%) | 12 (24%) | 16 (32%) | 16 (32%) | Hearing Loss | Yes <sup>15</sup> |
| Increasing social activity | 0 (0%) | 1 (2%) | 4 (8%) | 17 (37%) | 27 (53%) | Social Engagement | Yes <sup>15</sup> |
| Cognitive training | 0 (0%) | 2 (4%) | 10 (20%) | 20 (40%) | 18 (36%) | Cognitive Engagement | Some <sup>15</sup> |
| Following a Mediterranean diet | 1 (2%) | 4 (8%) | 16 (31%) | 22 (43%) | 8 (16%) | Mediterranean diet | Some <sup>16</sup> |
| Avoiding living near busy roads | 13 (25%) | 13 (25%) | 23 (45%) | 2 (4%) | 0 (0%) | Air Pollution | Some <sup>17</sup> |
| Doing crosswords or sudoku puzzles | 1 (2%) | 1 (2%) | 14 (27%) | 18 (35%) | 17 (33%) | Cognitive Engagement | Some <sup>15</sup> |
| Learning a new language | 1 (2%) | 0 (0%) | 11 (22%) | 27 (53%) | 12 (24%) | Cognitive Engagement/<br>Bilingualism | Some <sup>15</sup> |
| Following a DASH (Dietary Approaches to Stop Hypertension) diet | 3 (6%) | 1 (2%) | 21 (41%) | 18 (35%) | 8 (16%) | DASH diet | Some <sup>18</sup> |
| Drinking coffee | 2 (4%) | 11 (22%) | 26 (51%) | 12 (24%) | 0 (0%) | Coffee | Some <sup>15</sup> |
| Avoiding exposure to aluminium | 7 (14%) | 8 (16%) | 24 (47%) | 8 (16%) | 4 (8%) | Aluminium | Some <sup>19</sup> |
| Taking multivitamins | 9 (18%) | 11 (22%) | 21 (43%) | 7 (14%) | 1 (2%) | Vitamins | Some<sup>†</sup> <sup>20-22</sup> |
| Avoiding living near powerlines | 19 (37%) | 9 (18%) | 19 (37%) | 3 (6%) | 1 (2%) | Powerlines | None |
| Including coconut oil in their diet | 18 (35%) | 17 (20%) | 22 (43%) | 1 (2%) | 0 (0%) | Coconut oil | NA<sup>‡</sup> |
| Including turmeric in their diet | 17 (34%) | 7 (14%) | 19 (38%) | 7 (14%) | 0 (0%) | Curcumin | NA<sup>‡</sup> |
| Including ginkgo biloba in their diet | 14 (27%) | 8 (16%) | 29 (57%) | 0 (0%) | 0 (0%) | Ginkgo biloba | NA<sup>‡</sup> |

Notes. Responses are presented as N (%). N = total number of people who chose this response, % = proportion of participants who chose this response.
\* Alcohol shows a U-shaped curve with dementia where low alcohol consumption has been associated with better cognitive outcomes <sup>23)</sup>
† No reviews have been identified looking at multivitamin use specifically, however reviews have been conducted for individual vitamins
‡ No systematic reviews have been identified for this risk factor in the prevention of dementia

### Primary care provider roles in dementia prevention

The majority of participants agreed that general/family practitioners and practice nurses should be discussing dementia prevention with patients prior to age 65 (88%), however when conducting 45-49 year old chronic disease risk assessments, most participants responded that they only occasionally (sometimes, rarely or never) discuss dementia prevention (55%). Additionally, while the majority of participants frequently (often, very often or nearly always) assessed cardiovascular risk factors (78%) or discussed lifestyle changes (69%), they only occasionally (sometimes, rarely or never) discussed dementia prevention during these sessions (84%). However, the majority of participants agreed that it is: i) part of their role to discuss dementia prevention with their adult patients (78%), and ii) they should be discussing dementia prevention with more of their adult patients (76%). See Table 5.

**Table 5.**
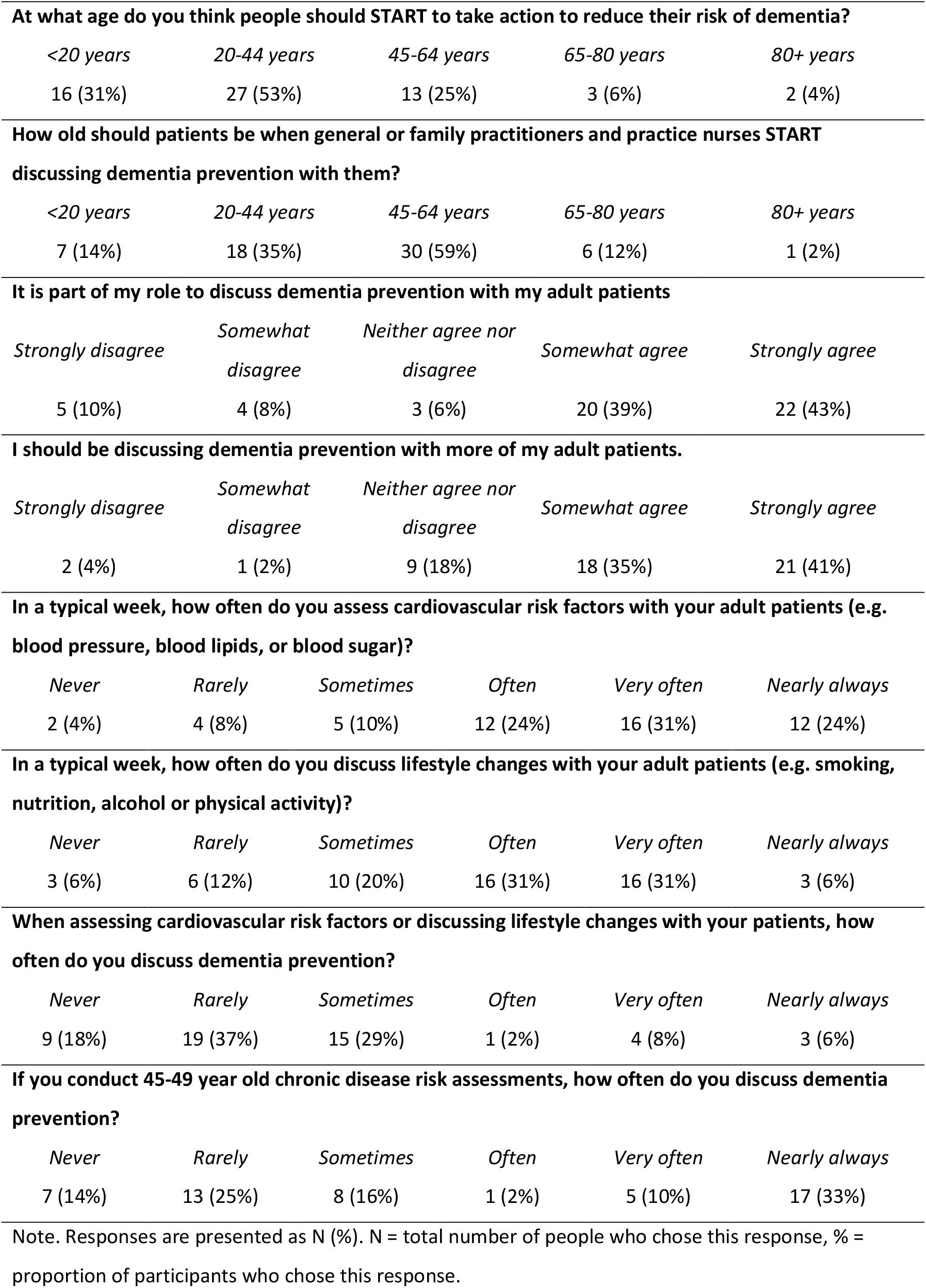
Responses to Questions about the Primary Healthcare Provider’s Role in Dementia Prevention

| *<20 years* | *20-44 years* | *45-64 years* | *65-80 years* | *80+ years* |
| --- | --- | --- | --- | --- |
| 16 (31%) | 27 (53%) | 13 (25%) | 3 (6%) | 2 (4%) |

| *<20 years* | *20-44 years* | *45-64 years* | *65-80 years* | *80+ years* |
| --- | --- | --- | --- | --- |
| 7 (14%) | 18 (35%) | 30 (59%) | 6 (12%) | 1 (2%) |

**It is part of my role to discuss dementia prevention with my adult patients**
| *Strongly disagree* | *Somewhat disagree* | *Neither agree nor disagree* | *Somewhat agree* | *Strongly agree* |
| --- | --- | --- | --- | --- |
| 5 (10%) | 4 (8%) | 3 (6%) | 20 (39%) | 22 (43%) |

**I should be discussing dementia prevention with more of my adult patients.**
| *Strongly disagree* | *Somewhat disagree* | *Neither agree nor disagree* | *Somewhat agree* | *Strongly agree* |
| --- | --- | --- | --- | --- |
| 2 (4%) | 1 (2%) | 9 (18%) | 18 (35%) | 21 (41%) |

| *Never* | *Rarely* | *Sometimes* | *Often* | *Very often* | *Nearly always* |
| --- | --- | --- | --- | --- | --- |
| 2 (4%) | 4 (8%) | 5 (10%) | 12 (24%) | 16 (31%) | 12 (24%) |

| *Never* | *Rarely* | *Sometimes* | *Often* | *Very often* | *Nearly always* |
| --- | --- | --- | --- | --- | --- |
| 3 (6%) | 6 (12%) | 10 (20%) | 16 (31%) | 16 (31%) | 3 (6%) |

| *Never* | *Rarely* | *Sometimes* | *Often* | *Very often* | *Nearly always* |
| --- | --- | --- | --- | --- | --- |
| 9 (18%) | 19 (37%) | 15 (29%) | 1 (2%) | 4 (8%) | 3 (6%) |

**If you conduct 45-49 year old chronic disease risk assessments, how often do you discuss dementia prevention?**
| *Never* | *Rarely* | *Sometimes* | *Often* | *Very often* | *Nearly always* |
| --- | --- | --- | --- | --- | --- |
| 7 (14%) | 13 (25%) | 8 (16%) | 1 (2%) | 5 (10%) | 17 (33%) |

Note. Responses are presented as N (%). N = total number of people who chose this response, % = proportion of participants who chose this response.

### Barriers and enablers to working with patients to reduce dementia risk

A total of 169 responses to barriers and 128 responses to recommendations were received (see Appendix 2 and 3 for a comprehensive list of responses). Thematic analysis indicated that, from the perspectives of primary healthcare providers, the most frequently reported barrier to working with patients to reduce dementia risk was that patients were not motivated to hear about dementia prevention. This included subthemes such as: the patient doesn’t understand dementia and/or its risk factors (e.g. “Client doesn’t understand what dementia is”, “Patients not believing they are at risk of dementia”); and patients don’t care or don’t want to talk about dementia and/or dementia risk reduction (e.g. “Many clients do not want to discuss the subject”, “Other issues prioritised by patients”). The second most frequently reported barrier was related to limitations of the current work structure or the structure of the health system. This included subthemes such as: no time or capacity to integrate into their appointments (e.g. “No time in consultation that usually focused around more acute needs”); and preventative health is not considered a priority by Medicare or by patients as the current culture is more focused on cure (e.g. “people’s value is in cure and not prevention”). The most common recommendation was to increase resources (e.g., brochures; clear guidelines on dementia risk reduction), followed by increasing awareness about dementia and positive messaging (e.g., TV advertising campaigns; reducing stigma around dementia). See Table 6 for a summary of the most commonly identified barriers and enablers.

**Table 6.** Commonly reported barriers and enablers to working with patients to reduce dementia risk. Ranked in order of the proportion of responses that related to the theme.

| Barriers | N | Proportion of responses related to this theme |
| --- | --- | --- |
| • Patient doesn't ask/listen | 48 | 28% |
| • Current work structure | 34 | 20% |
| • Lack of knowledge/ education/ resources for GP | 28 | 17% |
| • Patients unwilling to change current behaviour | 20 | 12% |
| • Medical appointments are not the right setting for this discussion | 14 | 8% |
| • Dementia prevention is too late for the patient | 11 | 7% |
| Enablers |  |  |
| • Increase resources | 49 | 38% |
| • Improving dementia awareness and messaging | 18 | 14% |
| • Increase dementia and dementia risk reduction education for primary healthcare staff | 16 | 13% |
| • Increase training for primary healthcare staff | 13 | 10% |
| • Change in policies or restructuring of health/Medicare system | 12 | 9% |

Note. Only response themes with more than 10 relevant responses are included in the table.

## Discussion

### Knowledge and perceptions about dementia risk reduction in primary care

Overall, the perceptions of participants closely matched the evidence in the literature for dementia risk factors suggesting that primary healthcare providers have broadly accurate knowledge about the major modifiable risk factors for dementia. However, continued efforts still need to be made to educate primary healthcare providers on the value of dementia risk reduction as the results showed that one third of primary healthcare providers do not agree that it is possible to reduce the risk of a person developing dementia. More training (e.g., prerequisite training during the early years of study or practice) may be needed as the majority of participants in this study (62%) reported that they had not received any training in dementia prevention.

PNs were found to have much less confidence in their knowledge of dementia risk factors than GPs. More research is needed to investigate the underlying reasons for this disparity. The results showed that the proportion of PNs that had received training in dementia risk factors was similar to that of GPs, however as only 6 PNs and 4 GPs had received any training on dementia risk factors, these results are difficult to interpret. It is likely that training and other support that is effective in raising confidence and self-efficacy might help to increase both factual and perceived knowledge of dementia risk factors and lead to a change in beliefs and practices.

### Barriers and enablers

While the majority of participants frequently assess dementia risk factors in their adult patients, they do not commonly discuss dementia prevention during these sessions. Thematic analysis of barriers to working with patients to reduce dementia risk indicated that this may be due to both patient (e.g. “lack of interest from patients [to discuss dementia risk reduction]”) and system level factors (e.g. “preventative health not considered a priority by Medicare”). Interestingly, while most practitioners responded that people should start to take action to reduce their risk of dementia early (20–44 years old), they also responded that they shouldn’t discuss dementia prevention with patients until much later (45–64 years old). This may be because the aforementioned barriers are even more pertinent with younger patients (e.g. “[patients have a] lack of motivation for something that might not happen”). Multi-level changes are likely needed to overcome these barriers, including changes at the policy and system level (e.g., to help encourage and promote a culture of preventative care with financial incentives) and at the societal level (e.g., to improve dementia risk reduction awareness and advocacy among the general public).

Some of the recommendations suggested by healthcare professionals are achievable with collaborative efforts from researchers, developers and the media (e.g., increasing resources and increasing awareness and education of existing resources and information within the clinic setting). Many resources have been, or are currently in the process of being developed (e.g., ^11^). However, previous research has highlighted that there are difficulties faced with implementing effective educational resources and facilitating practice change in primary care ^12, 13^. Travers and colleagues conducted a review of educational and behavioural interventions for risk reduction in primary care and suggested that these approaches are at most, moderately effective ^12^. This is consistent with other work by Steiner and colleagues who also found that GPs were unsure of what dementia resources were available and how to access them ^14^. This indicates that resource creation is necessary, but not sufficient to generate practice change.

### Next steps

Previous research has surmised that the implementation of dementia risk reduction activities in primary care requires individualised knowledge translation and implementation strategies that are adapted to the specific barriers and needs of the relevant practice or organisation ^12, 13^. It has also been suggested that primary healthcare providers should be supported from an organisational level (to ensure the provision of adequate educational materials and training and as well as remunerated time for dementia prevention education) and that any knowledge material should have a clear presentation and purpose, use simple and concise language, be practical in its use and implementation, be readily accessible during a consultation and be based on sound and reliable evidence ^12^. The successful implementation of risk reduction activities into primary practice will therefore likely require a combination of carefully designed resources and appropriate and bespoke knowledge translation plans.

The next step in this research is to develop tailored tools and resources based on the recommendations described here and evaluate whether they are effective in improving risk reduction evidence uptake in primary care. The most frequently reported barrier in this study was that patients were not motivated to hear about dementia prevention advice during their medical consultations. Media campaigns aimed at raising awareness of dementia risk reduction activities across the lifespan may be helpful in this regard. Additionally, information sheets, brochures and risk assessment apps that are easily accessible within each medical practice may be beneficial in helping to educate both patients and clinicians. It may also be the case that primary healthcare providers are not fully aware of the number of people who want to hear dementia risk reduction advice. For example, Millard and colleagues ^9^ surveyed 621 patients over 30 years of age in GP waiting rooms and found that 78% of participants wanted to learn more about reducing their risk of dementia. In this circumstance, it may be beneficial to raise awareness of these statistics among primary healthcare providers. The second most frequently reported barrier - changes to current practice structures, will likely require top-down intervention from practice leads, regulators and policy makers. Effective knowledge translation plans that are developed collaboratively between researchers, clinicians and policy makers may be necessary to overcome this barrier.

### Limitations

These findings should be interpreted in context of some limitations. Firstly, the majority of respondents were female, indicating that male healthcare providers may have been under-represented in this sample. Secondly, as participation in the survey was voluntary, the respondents that completed the survey may represent a group of more well-informed and motivated healthcare providers. These findings need to be replicated in a larger sample of primary healthcare providers to understand how far the knowledge and views reported here can be generalised. Thirdly, there is a growing body of trial evidence on the effectiveness dementia risk reduction techniques which does not always coincide with the epidemiological evidence on dementia risk factors. This study did not separate primary healthcare providers’ knowledge of dementia risk factors from dementia risk reduction techniques (as the evidence base regarding the effectiveness of risk reduction techniques is still in its infancy). The separation of the two may be necessary in future studies of a similar nature once the evidence base grows more substantial.

## Conclusion

The findings from this study showed that Australian primary healthcare providers have accurate knowledge of the modifiable risk factors for dementia, however they face a number of patient, policy and system level barriers when working with patients to reduce dementia risk. Collaborative efforts between researchers, the media, clinicians and policy makers will likely be necessary to support the implementation of the identified recommendations into practice.

## Data Availability

The data are available on request

## Acknowledgments

We would like to acknowledge funding from the Dementia Centre for Research Collaboration and thank all the participants for their contributions to the study.

## Ethics Approval Statement

This study was approved by the UNSW HREC (HC3120)

